# How much pain is too much? Expectations of pain during intrauterine device insertion among Australian women: findings from an online survey

**DOI:** 10.64898/2026.05.21.26353829

**Authors:** Jacqueline Coombe, Jane L Goller, Helen Bittleston, Claire Felix-Faure, Henrietta Williams, Cassandra Caddy

## Abstract

There are several barriers to uptake of intrauterine devices (IUDs), with the fear of pain during insertion an emerging concern. Using data from an online survey, we sought to understand the experience of women who had undergone IUD insertion, with a particular focus on their expectation compared with their reported experience of pain. We found that, while most participants expected a moderate level of pain at insertion, many reported a high level of pain. Pain relief offered was variable, and, aside from that administered by an anaesthetist, no single method appeared to significantly reduce reported pain.

## Introduction

There are persistent barriers to the provision of intrauterine devices (IUDs) to Australian women. A lack of knowledge or misinformation about IUDs (Daniele, Cleland et al. 2017), a disinclination to use these devices at particular times of their reproductive lives (Coombe, Harris et al. 2018) and fear of pain during insertion (Potter, Rubin et al. 2014) can deter use. Challenges in accessing IUD insertion also include the availability of trained healthcare providers (Lodge, Sanci et al. 2017, Mazza, Watson et al. 2023).

Women often report IUD insertion as a painful experience, with a recent Australian study reporting that half of their sample (n=157/322) perceived pain during insertion as moderate or severe (Camões-Costa, Kwok et al. 2026). Of note, other studies have shown that women who have given birth vaginally typically report lower levels of pain (Allen, Carey et al. 2014). In addition, research shows that providers often underestimate the pain experienced during IUD insertion (Maguire, Morrell et al. 2014). There is mixed evidence regarding the most effective pain relief option to use during insertion (Nguyen, Lamarche et al. 2020). In Australia, guidelines in relation to IUD insertion include several pharmacological options to be used at the discretion of the provider (RANZCOG 2024, Royal Australian College of General Practitioners 2024). These include local anaesthetic cream applied to the cervix and cervical opening, local anaesthetic for para-or intracervical block, lidocaine spray applied to the cervical surface and os, or *Penthrox* (methoxyflurane) inhaler (Royal Australian College of General Practitioners 2024). Those undergoing IUD insertion in certain settings may also have the procedure undertaken under general anaesthetic.

Despite efforts to increase IUD uptake in Australia, we know little about the experiences of Australian women undergoing IUD insertion, and in particular their expectations of pain, and the relationship of this with the pain they experience during insertion. Concerningly, recent qualitative research found negative flow-on effects among a small group of women who reported experiencing pain beyond their expectations, including impacts on intimate relationships and willingness to undergo other gynaecological procedures (Caddy, Temple-Smith et al. 2025). This work highlights the importance of understanding and addressing experiences of pain during IUD insertions. As part of a broader online survey investigating the IUD insertion experience of Australian women (Felix-Faure, Coombe et al. 2025), this study explored the data relating to expected and actual pain experienced during insertion.

## Methods

### Survey design and study participants

We administered an online survey using Qualtrics software for 6 weeks from May to July 2023. Women and people with female reproductive organs, aged 18-45 years and living in Australia were eligible to participate if they had an IUD inserted (successful or unsuccessful) in the past two years. Taking approximately 20 minutes to complete, the survey comprised largely qualitative questions to investigate the experience of IUD insertion with several closed questions to collect contextual information. The survey also asked participants to rate the level of pain: a) expected and b) actually experienced during insertion on a numerical scale of 1-10, whereby a score of 1 represented no pain and a score of 10 represented the worst pain imaginable. This manuscript reports the findings for the numerical pain-rating. Our qualitative findings are reported elsewhere (Felix-Faure, Coombe et al. 2025).

Potential survey participants were people who had completed a previous study with the research team and consented to being recontacted regarding related research. These people received an email explaining the study, including a plain language statement, and link to the survey. A survey flyer was also circulated among researchers’ social media networks (e.g. professional X account or personal social media). Participants were not offered an incentive for survey completion. All participants were required to consent to participate before commencing the survey. This study was approved by the University of Melbourne Ethics Committee (ID:23972).

### Data management and analysis

Only data for participants who responded to the numerical pain-rating scale items were included. Consistent with another study (Akintomide, Nataliya et al. 2015), we created a categorical variable of severe (7 to 10) / non-severe (1 to 6) pain. We also created categorical variables for age group (18-29 years, 30-45 years); having given birth vaginally prior to IUD insertion (yes / no); IUD type (copper or hormonal / excluding those reporting copper and hormonal); IUD inserter (GP / specialist) and reason for IUD insertion (non-gynaecological / gynaecological reasons). We also created a variable for pain relief at insertion categorised as: i) no pharmacological pain relief, ii) medication taken prior to appointment (e.g. over counter pain tablets, prescription medication), iii) pain relief provided by healthcare provider during insertion (e.g. lidocaine, *Penthrox*, nitrous oxide) and, iv) by anaesthetist or an accompanying HCP other than the inserter and includes general anaesthetic and those who commented they were provided a spinal, epidural or sedation such as fentanyl or medazolam. Those who selected multiple types of pain relief were included in whichever category had the potential for the highest amount of pain relief (for example, if they selected over the counter pain tablets and lidocaine, they were put into category iii) being pain relief provided by healthcare provider during insertion). We explored characteristics associated with the outcome ‘severe’ pain experienced using univariable and multivariable logistic regression. All variables associated with severe pain in univariable models were retained in the final multivariable model, with the exception of pain relief, where a decision was made *a priori* to retain this due to its potential impact on the outcome. Analyses were conducted using Stata V.18.0.

## Results

Of the total of 220 eligible people who consented to participate in the survey, 81.4% (n=179) answered numerical pain-rating questions and formed the study population.

Most participants (94.4%, n=169) were cisgender women, 5% (n=9) were trans and gender diverse and one preferred not to say. Two-thirds (65.9%, n=118) reported their sexuality as heterosexual, 50.8% (n=91) were aged 18-29 years and 20.7% (n=37) had given birth vaginally prior to IUD insertion. The majority (>97%) were permanent residents of Australia and Medicare card holders. Most participants (84.9%, n=152) had a hormonal IUD with Mirena (n=136) the most common; 9.5% (n=17) had a copper IUD and 5.6% (n=10) had used both a hormonal and a copper IUD. Over half (54.2%, n=97) had an IUD inserted for gynaecological reasons (for example, to manage endometriosis symptoms or painful/heavy periods). IUDs were most frequently inserted by a specialist health care provider (60.3%, n=108) followed by a GP (28.5%, n=51), with 11.2% (n=20) reporting that they had an IUD inserted by a GP and a specialist. Pharmacological pain relief reported on prior to or during IUD insertion included none (14.3%, n=25), medication taken prior to the appointment (22.9%, n=40), pain relief provided during insertion (38.9%, n=66) and sedation or general anaesthetic (24.0%, n=44).

The level of pain expected and experienced during IUD insertion is shown in Figure 1. On the pain-rating scale of 1 to 10, the median pain level expected was 5 (interquartile range (IQR) 4-7), and experienced was 7 (IQR 3-9). Just over half of participants (n=96, 53.6%) experienced severe pain during IUD insertion.

**Figure 1.**
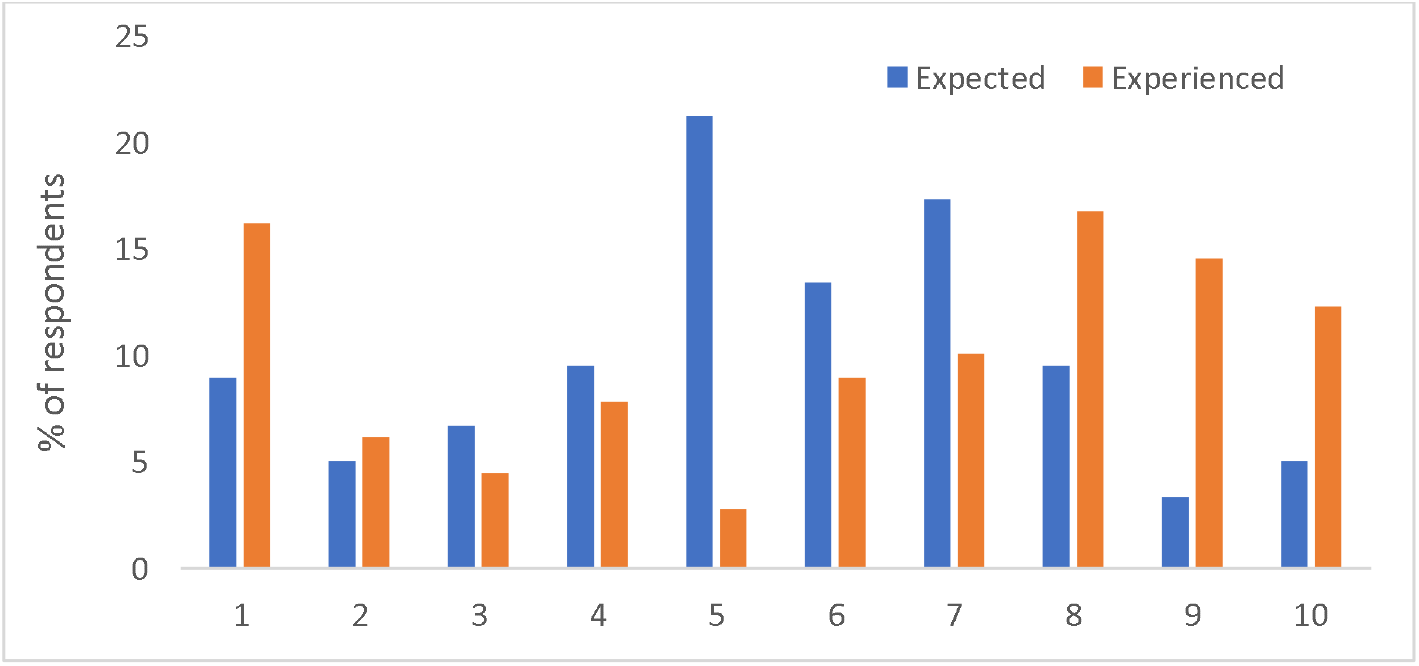
Distribution of expected and experienced pain scores, n=179.

The univariable and multivariable results are provided in Table 1. Adjusting for age-group, IUD inserter type, pain relief and having given birth vaginally, multivariable analysis showed that those less likely to experience severe pain had given birth vaginally prior to IUD insertion (AOR 0.22, 95%CI 0.07-0.68) or had received a general anaesthetic or sedation at insertion (AOR 0.26, 95%CI 0.07-0.95).

**Table 1.**
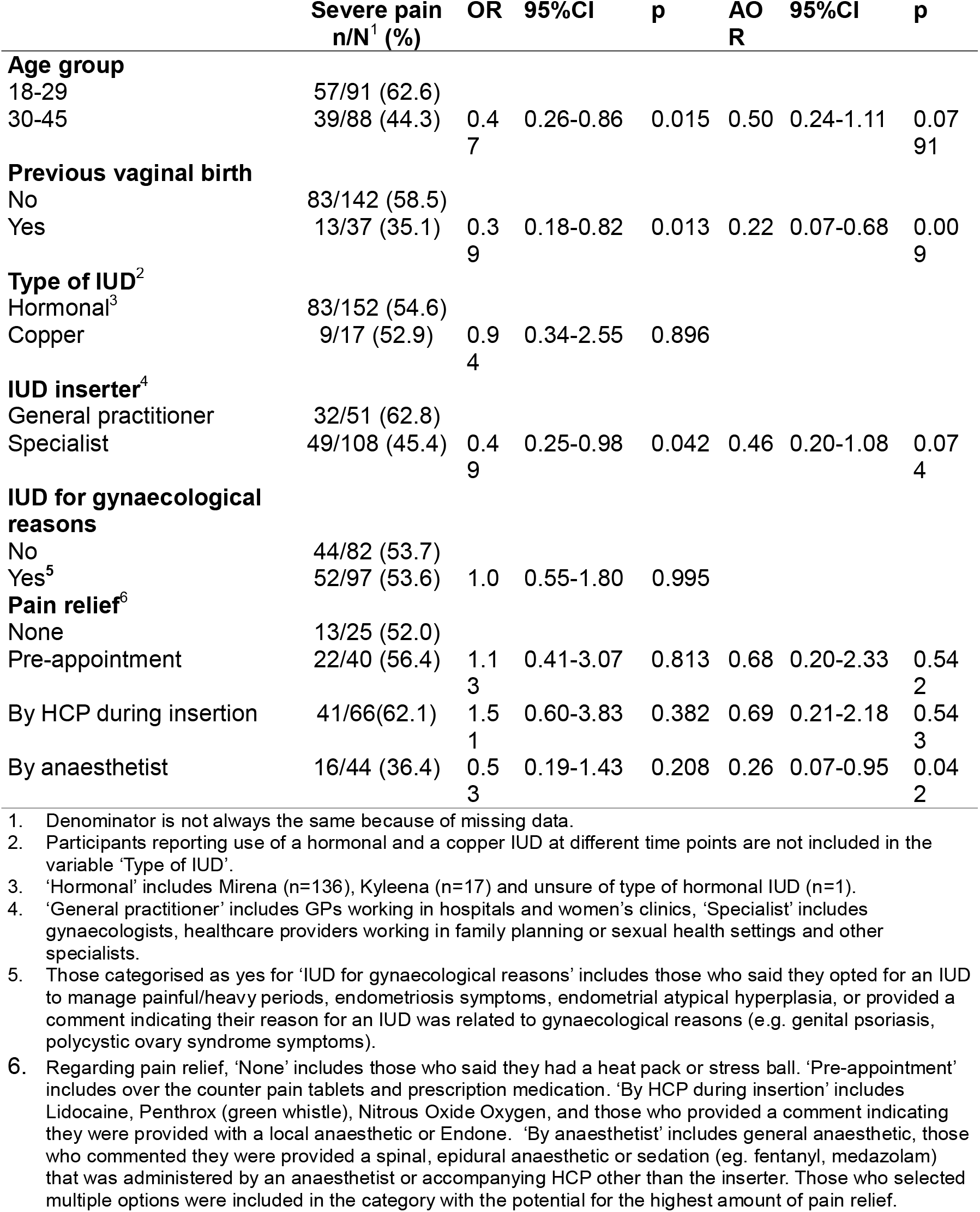
Associations with severe pain experienced on IUD insertion.

|  | Severe pain<br>n/N<sup>1</sup> (%) | OR | 95%CI | p | AO<br>R | 95%CI | p |
| --- | --- | --- | --- | --- | --- | --- | --- |
| **Age group** |  |  |  |  |  |  |  |
| 18-29 | 57/91 (62.6) |  |  |  |  |  |  |
| 30-45 | 39/88 (44.3) | 0.47 | 0.26-0.86 | 0.015 | 0.50 | 0.24-1.11 | 0.0791 |
| **Previous vaginal birth** |  |  |  |  |  |  |  |
| No | 83/142 (58.5) |  |  |  |  |  |  |
| Yes | 13/37 (35.1) | 0.39 | 0.18-0.82 | 0.013 | 0.22 | 0.07-0.68 | 0.009 |
| **Type of IUD<sup>2</sup>** |  |  |  |  |  |  |  |
| Hormonal<sup>3</sup> | 83/152 (54.6) |  |  |  |  |  |  |
| Copper | 9/17 (52.9) | 0.94 | 0.34-2.55 | 0.896 |  |  |  |
| **IUD inserter<sup>4</sup>** |  |  |  |  |  |  |  |
| General practitioner | 32/51 (62.8) |  |  |  |  |  |  |
| Specialist | 49/108 (45.4) | 0.49 | 0.25-0.98 | 0.042 | 0.46 | 0.20-1.08 | 0.074 |
| **IUD for gynaecological reasons** |  |  |  |  |  |  |  |
| No | 44/82 (53.7) |  |  |  |  |  |  |
| Yes<sup>5</sup> | 52/97 (53.6) | 1.0 | 0.55-1.80 | 0.995 |  |  |  |
| **Pain relief<sup>6</sup>** |  |  |  |  |  |  |  |
| None | 13/25 (52.0) |  |  |  |  |  |  |
| Pre-appointment | 22/40 (56.4) | 1.13 | 0.41-3.07 | 0.813 | 0.68 | 0.20-2.33 | 0.542 |
| By HCP during insertion | 41/66(62.1) | 1.51 | 0.60-3.83 | 0.382 | 0.69 | 0.21-2.18 | 0.543 |
| By anaesthetist | 16/44 (36.4) | 0.53 | 0.19-1.43 | 0.208 | 0.26 | 0.07-0.95 | 0.042 |

1. Denominator is not always the same because of missing data.
2. Participants reporting use of a hormonal and a copper IUD at different time points are not included in the variable 'Type of IUD'.
3. 'Hormonal' includes Mirena (n=136), Kyleena (n=17) and unsure of type of hormonal IUD (n=1).
4. 'General practitioner' includes GPs working in hospitals and women's clinics, 'Specialist' includes gynaecologists, healthcare providers working in family planning or sexual health settings and other specialists.
5. Those categorised as yes for 'IUD for gynaecological reasons' includes those who said they opted for an IUD to manage painful/heavy periods, endometriosis symptoms, endometrial atypical hyperplasia, or provided a comment indicating their reason for an IUD was related to gynaecological reasons (e.g. genital psoriasis, polycystic ovary syndrome symptoms).
6. Regarding pain relief, 'None' includes those who said they had a heat pack or stress ball. 'Pre-appointment' includes over the counter pain tablets and prescription medication. 'By HCP during insertion' includes Lidocaine, Pentrox (green whistle), Nitrous Oxide Oxygen, and those who provided a comment indicating they were provided with a local anaesthetic or Endone. 'By anaesthetist' includes general anaesthetic, those who commented they were provided a spinal, epidural anaesthetic or sedation (eg. fentanyl, medazolam) that was administered by an anaesthetist or accompanying HCP other than the inserter. Those who selected multiple options were included in the category with the potential for the highest amount of pain relief.

## Discussion

In our study, a history of vaginal birth was the strongest predictor of the pain experienced during IUD insertion, with these women five-fold *less likely* to experience severe pain. This finding is consistent with previous research (Lopes-Garcia, Carmona et al. 2023), and highlights the importance of discussing history of birth during a pre-IUD insertion consult in regards to potential experience of pain. Although other studies such as Camoes-Costa et al (Camões-Costa, Kwok et al. 2026) report a history of pregnancy as important in the perception of pain during IUD insertion, we note that these did not collect data on history of birth (vaginal or caesarean). While women having an IUD inserted under general anaesthetic or sedation were also less likely to report severe pain, we found no other relationship between type of pain medication, provider or type of IUD and reported pain.

While most of our participants expected a moderate level of pain at IUD insertion, many reported a high level of pain. This contrasts with previous reports that those who *expect* a high level of pain will also *experience* a high level of pain (Allen, Carey et al. 2014). This has in part been attributed to pre-procedural anxiety and these studies have supported anxiety reducing interventions to improve experience (Dina, Peipert et al. 2018, Akdemir and Karadeniz 2019). However, our findings show that even with moderate expectations of pain prior to insertion, over half of participants experienced severe pain. Our findings therefore caution over emphasis on managing pre-procedural anxiety and instead providing patients with information about expectations during the procedure, including pain relief options available.

Only those who had their IUD inserted under a general anaesthetic or sedation were less likely to experience severe pain. Surprisingly, other pain relief methods did not have a significant impact on actual experiences of pain in our study. Our findings also showed high variability in the type of pain medication provided to participants. This is consistent with variable evidence in the literature on the best pharmacological strategies to manage pain during IUD insertion (Rahman, King et al. 2024), and recent work conducted with Victorian healthcare providers regarding their provision of pain relief during IUD insertion that demonstrated providers take a flexible approach to pain management based on the individual patient (Wood, Caddy et al. 2025). Our findings support further investigation into effective pharmacological pain management strategies for IUD insertion. In the meantime, in line with the recommendations by Tarrant et al (Tarrant, Grills et al. 2025), examination of current clinical practice guidelines to ensure healthcare providers are supported and able to provide appropriate pain management options to their patients is warranted. Further exploration into the needs of women prior to, during and after IUD insertion is also needed. This includes a better understanding of the influence of pain expectation prior to insertion, as well as what information women are provided about pain prior to the procedure, the pain management options they are offered and how this impacts their experiences.

The limitations of our study include its small size, the potential impact of an up to two-year time frame between IUD insertion and our survey, and the exclusion of some participants from the final multivariable model. The latter was due to selecting multiple IUD types (i.e. we were interested in their most recent insertion, but we were unable to differentiate whether they were reporting on their hormonal or copper IUD insertion).

Our study showed that many women report significant pain during IUD insertion. Ensuring that all women undergoing IUD insertion are offered appropriate analgesia is vital and clinical trials investigating the optimal pain relief for IUD insertion are urgently needed. In the meantime, women should always be offered some form of pain relief, as recommended in current clinical guidelines.

## Data Availability

The participants of this study did not give written consent for their data to be shared publicly, so due to the sensitive nature of the research supporting data is not available.

## Notes

### Competing Interest Statement

The authors have declared no competing interest.

### Author Declarations

This study was approved by the University of Melbourne Ethics Committee (ID:23972).

